# Prevalence and factors associated with help-seeking behavior among female youth who experienced Intimate Partner Violence in Tanzania, 2015/16 and 2022

**DOI:** 10.1101/2025.10.29.25339112

**Authors:** Asteria Karungi Nyongoli, Idd A. Muruke, Martha Reuben Magili, Anthony Charles Kavindi, Laura J. Shirima, Winfrida C. Mwita, Caroline Amour¹, Sia E. Msuya

## Abstract

**Background:** Intimate Partner Violence (IPV) remains a significant global public health concern, with youths disproportionately affected compared to the general population of women aged 15-49 years. In Tanzania, nearly one in two (47.8%) female youth reported having experienced IPV in 2015. Despite national interventions, help-seeking behavior among this group remains low.

**Methods:** The study was an analytical cross-sectional study design, utilizing data from the 2015/16 and 2022 Tanzania Demographic and Health Surveys (TDHS). Descriptive statistics were used to estimate the prevalence of help-seeking behavior and sources of help used, while Modified Poisson regression was used to determine factors associated with help-seeking behavior among female youth who experienced IPV within the 12 months preceding the survey. Statistical significance was set at a p-value of ≤ 0.05.

**Results:** Findings from this study showed a significant decline in help-seeking behavior from 52.0% in 2015/16 to 34.1% in 2022 (p-value <0.001). Most youth sought help from informal sources, such as their own or the partner’s family, while formal sources like medical personnel, police and social welfare organizations were among the least utilized. Help-seeking behavior was significantly associated with belonging to the rich wealth index (aPR= 1.76; 95%CI: 1.05- 2.97), working (aPR= 1.82; 95%CI: 1.15-2.88) and being younger than the spouse.

**Conclusion:** These findings highlight the need to strengthen existing programs and interventions by promoting family engagement in IPV awareness and response efforts given the critical role families play in early detection, emotional support and referral services. Interventions should also aim to address cultural norms that may discourage seeking help outside the family, particularly those rooted in stigma and fear of judgement. Youth who are not in school should be empowered economically so as to reduce their dependency on intimate partners. Increased economic autonomy may enhance their ability and willingness to seek help in cases of violence, as financial reliance often limits options for escape and recovery.

## Introduction

Intimate partner violence is the most common form of Gender Based Violence and remains a significant global public health concern. Worldwide, one in every three women is subjected to physical and/or sexual abuse in their lifetime, most often perpetrated by an intimate partner (1). This reflects the extent of gender inequality and discrimination against women. Youth represents a critical developmental stage characterized by rapid physical, emotional and social changes (2). Evidence suggests that female youth experience a disproportionately higher prevalence of IPV compared to the general population of women aged 15-49 years. In Tanzania, nearly one in two ( 47.8%) female youth aged 15-24years reported having experienced IPV as of 2015 (3) while IPV in the general population was 41.6% (4). Help-seeking behavior is crucial to overcome the short and long term consequences of IPV (5).

Help-seeking behavior among female youth who experienced IPV in Sub-Saharan Africa is notably low. Only 31.0% of adolescent girls aged 15-19 years and 35.6% of those aged 20-24 years sought help (6). In Kenya, the prevalence is even lower at 21.7% among female youth aged 15-24 years (7). In Tanzania, despite the documented higher prevalence of IPV among female youth, there is limited study specifically reporting the prevalence of help-seeking behavior in this group.

Not seeking help after experiencing IPV is associated with increased feelings of isolation, shame and, depression, often leading to mental health problems. The absence of support may also result in chronic stress and anxiety, which can manifest in harmful coping mechanisms such as substance abuse. Furthermore, not seeking help can perpetuate ongoing cycles of violence and suffering (6,8–10).

To address this, the World Health Assembly adopted a global plan of action aimed at strengthening health systems as part of a multisectoral response to violence, particularly against women, girls and children (11). Tanzania was one of the four lead countries in the Global Partnership to End Violence, which emphasized the action needed to prevent and respond to violence.

Through the National Plan of Action to End Violence Against Women and Children, numerous interventions have been implemented nationally to encourage reporting and ultimately reduce IPV. These include peer and community support groups such as “girl clubs” and “My choice, My right” as well as UNFPA led programs. Community level committees like “MTAKUA”. Community awareness campaigns and dialogues targeting influential community leaders have been established, alongside capacity-building initiatives for the frontline workers including social welfare officers, police and health care personnel. Additional measures include ensuring the presence of the Police Gender and Children’s Desk across the country, the expansion of One Stop Centers offering medical treatment, psychosocial support, counseling, and legal assistance to survivors. A toll-free telephone outreach service for children accessible across all networks. Health facilities have also been equipped with GBV management guidelines applicable at all levels of care. The National Plan of Action aimed to reduce all forms of violence by 50% and to ensure that 65% of survivors report the incident within 72 hours by 2022 (11).

Although the 2022 TDHS indicates a decline in all forms of IPV from the reported prevalences in 2015/15, the prevalence rates remain above the targets set by the 2017/18- 2021/22 NPA-VAWC. In 2022, the overall IPV in the last 12 months preceding the survey was 36.8% compared to 44.8% in 2015/16. Between the 2015/16 and 2022 TDHS, physical IPV in the past 12 months declined from 29.9% to 27.6%, sexual IPV from 12.4% to 11.8% and emotional IPV from 33.5% to 23.7%. Despite the interventions in place, there is a concerning increase in the proportion of women who do not seek help after experiencing IPV from 54% in 2015/16 to 38% in 2022 (11–13).

These findings suggest that, despite the high prevalence of IPV and the wide range of interventions in place, a substantial proportion of women particularly youth do not seek help following IPV (8,13,14). This highlights a critical gap in the existing response. Therefore, this study aims to look at the prevalence of help-seeking behavior among female youth who experienced IPV and to identify the factors associated with this behavior.

## Materials and Methods

### Data source

The study analyzed data from two rounds of the recent Tanzania Demographic and Health Surveys of 2015/16 and 2022 conducted across all the regions of the country. The sampling technique used was a two-stage cluster sampling. The sampling frame consisted of a list of all enumeration areas (EAs) identified in the previous census. Both the 2015/16 and 2022 TDHS used EAs based on the 2012 Tanzania Population and Housing Census. In the first stage, clusters (EAs) were selected using probability proportional to size, 608 EAs in the 2015/16 and 629 in 2022. In the second stage, households were randomly selected within each EA. All women aged 15-49 who were either the usual residents or visitors the night before the survey were eligible for interview. A male survey was also conducted in a subsample of households, every third household in 2015/16 and every second household in 2022. Additionally, the domestic violence module was administered to one randomly selected woman per household within the male subsample households in the 2022 survey. Information on domestic violence was obtained through the Woman’s questionnaire.

### Study design and sample size

The current study employed analytical cross-sectional study design. The total sample included 954 female youth aged 15-24 years who participated in the domestic violence module and reported experiencing intimate partner violence within the 12 months preceding the surveys, 681 from the 2015/16 and 273 from the 2022 TDHS (Fig 1).

**Fig 1.**
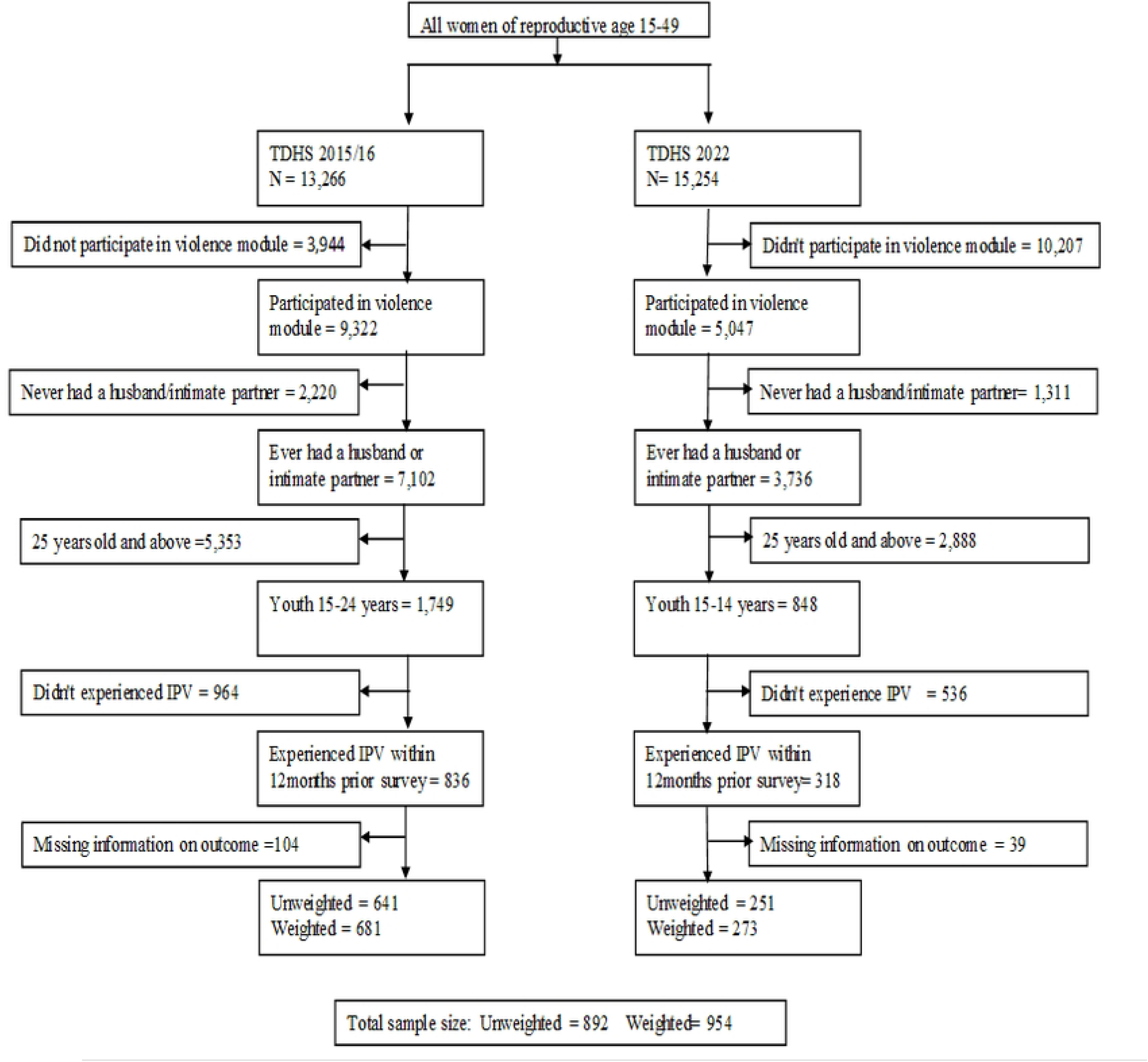
Flow Chart for selection of study participants

### Study Variables

#### Dependent variable

The dependent variable in this study was Help-seeking behavior following intimate partner violence. It is binary variable, coded “0” for no help was sought and “1” for sought help from someone. Data on help-seeking was derived from the survey question: “*Thinking about what you yourself have experienced among the different things we have been talking about, have you ever tried to seek help to stop the person(s) from doing this to you again? (*Response options: *yes/no)”*.

Potential sources of help included both informal and formal sources. Informal sources included the woman’s own family, her partner’s or husband’s family, friends, neighbors and current or former boyfriends. Formal sources included the police, criminal justice system, health care staff, social services, religious leaders.

#### Independent variables

The independent variables in this study included the respondent’s socio-demographic characteristics, partner-related characteristics, characteristics of the violence experienced, attitudes towards wife beating and women empowerment indicators.

Respondent’s age and partner age were collected as continuous variable. Spousal age difference was measured in years, obtained by subtracting respondent’s age from partner’s age. Respondent’s age was categorized into two categories, adolescent girls (15–19) and young women (20–24). Spousal age difference and number of living children were categorized based on literature. Wealth index, fear of the partner, education level of the participant and that of the partner were recategorization guided by related previous literatures.

Variables; residence, geographical zone, working status, partner’s alcohol intake, history of witnessing interparental violence and experience of severe violence were collected as categories and were used as how they were collected.

Justifying wife beating was a composite variable from five questions which were asked to the respondent that, “wife beating is justified if; (i) wife goes out without telling the husband (ii) wife neglects the children (iii) wife argues with the husband (iv) wife refuses to have sex with the husband (v) wife burns the food”. Each of the five questions required a yes or no or I don’t know response. In this study respondent was considered to justify wife beating if she responded yes to any of the five questions and otherwise was considered not to justify wife beating. Participation in decision making was also a composite variable derived from three questions asking the person who usually decides on; (i)respondent’s health care (ii)large household purchase (iii)visits to family or relative. The three questions had responses; respondent alone, respondent and husband/partner, husband/partner alone and someone else. In this study, female youth was considered to participate in decision making if she responded “respondent alone” or “respondent and husband/partner” (Table 1).

**Table 1.**
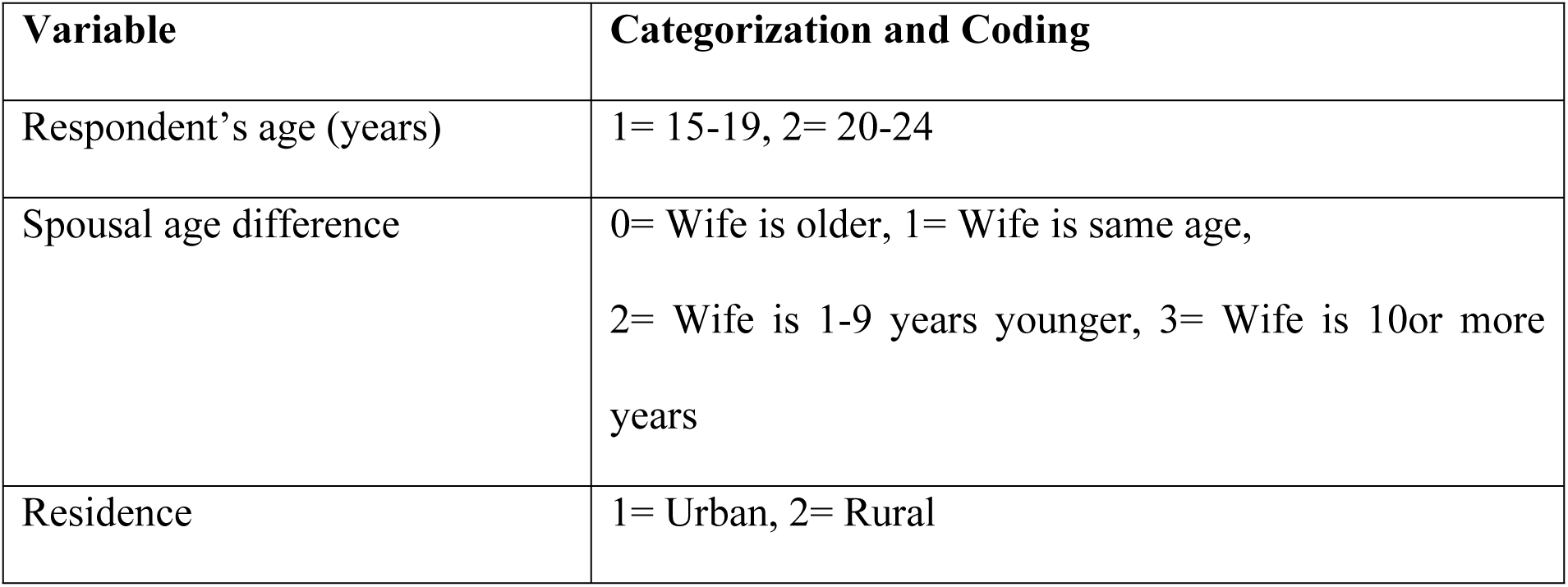
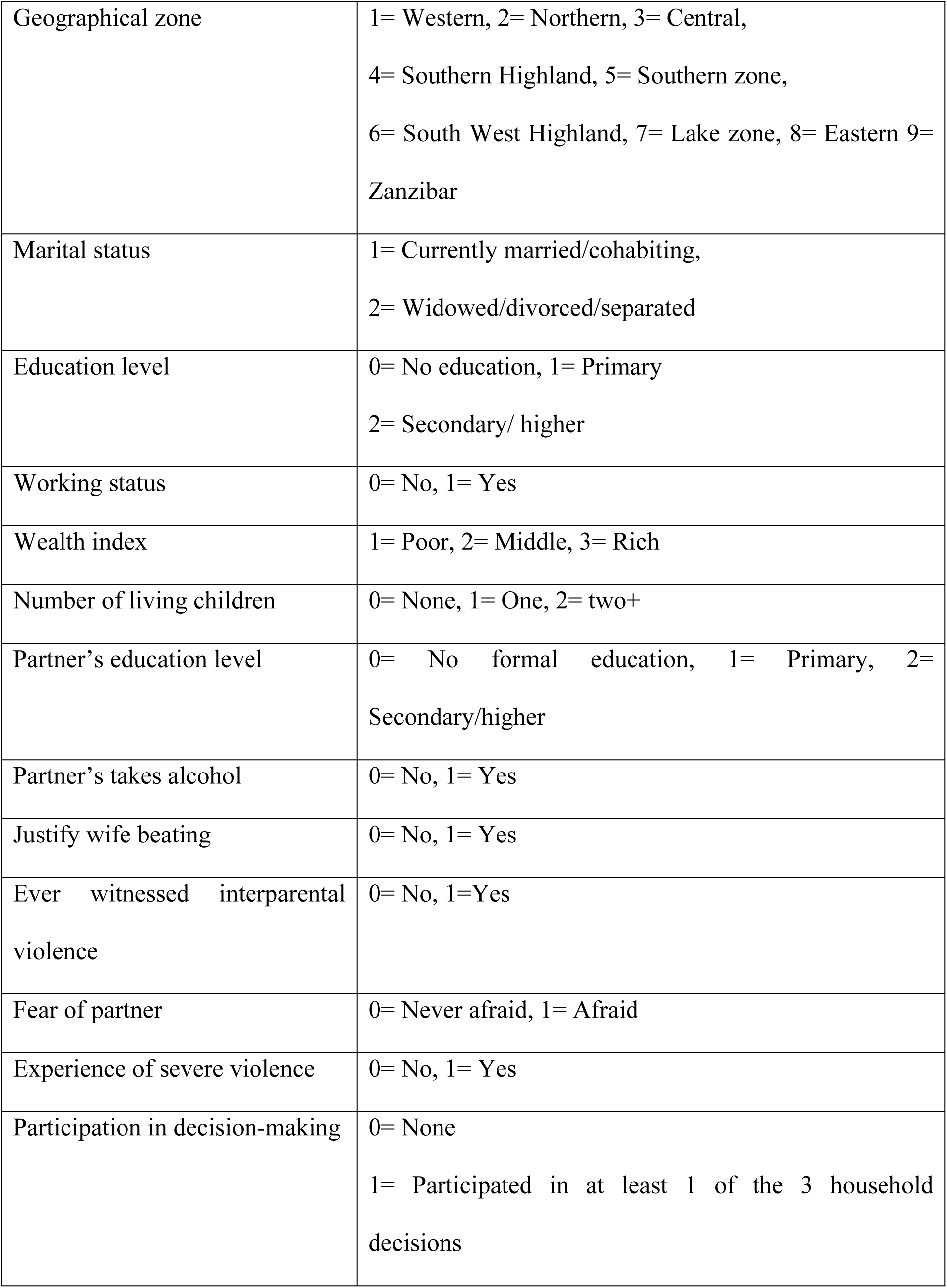
Categorization and coding of independent variables.

### Statistical analysis

Data cleaning and analysis was conducted using STATA software version 17.0. Preliminary data exploration was carried out to ensure the availability of key variables, assess missing values and ensure consistency in variable names across the two survey years. Respondents with missing values on the outcome variable were excluded from the analysis. Variables; women participation in decision making, partner’s education level and spousal age difference had missing of less than 5% which was delt by doing complete case analysis. Some variables were re-categorized based on previous literatures to enhance interpretability and comparability across studies.

Analyses were conducted separately for the 2015/16 TDHS and 2022 TDHS datasets. To account for the complex survey design, all analyses were weighed and adjusted for clustering. A sub-population of female youth aged 15-24 years who reported experiencing Intimate partner violence and had complete data on the outcome variable was used for all analysis.

Descriptive statistics were presented as frequencies and percents for categorical variables. For continuous variables, measures of central tendency and corresponding measures of dispersion were used. Factors associated with help-seeking behavior were analyzed using 2022 dataset.

Classical logistic regression was initially considered to assess factors associated with help-seeking among female youth who experienced Intimate partner violence. However, it was discarded due to its tendency to overestimate prevalence ratios and confidence intervals when the outcome prevalence exceeds 10% (15,16), as was the case in both surveys. Therefore, other alternatives to logistic regression were considered.

Log binomial was then attempted but encountered convergence issues. Thus, Poisson regression with a log link and robust standard errors was used to estimate prevalence ratios and 95% confidence interval. The “svy” command was applied without specifying the “vce (robust)” option wasn’t added as it automatically incorporates robust variance estimation.

Both crude and adjusted analyses were performed separately for each survey to identify the factors associated with predictors and help-seeking behavior over time.

Variables that were significant at p-value less or equal to 0.05 in the crude Modified Poisson regression, as well as those previously reported in literatures as significant or important to influence help-seeking behavior were considered for multivariable regression analysis. The best fit model was selected based on Akaike Information Criteria (AIC) and Bayesian Information Criterion (BIC). The model with the lowest Akaike Information Criteria (AIC) and BIC was considered the most parsimonious. To assess multicollinearity among independent variables, Spearman correlation was performed and no significant correlation found, indicating no multicollinearity. Variables with p- value less or equal to 0.05 in the multivariable analysis were considered significantly associated with help-seeking behavior among female youth who experienced violence.

### Ethical consideration

The original DHS data collection followed ethical standards including informed consent and confidentiality. Ethical approval for this study was obtained from the KCMC University Clinical Research and Ethical Review Committee (KCMU-CRERC) under clearance number PG 165/2024 Permission to access and use the Tanzania Demographic and Health Survey data was granted by DHS MEASURE program. The data was used for the purposes of this academic research, in accordance with the terms of use.

## Results

### Respondents Background Characteristics

A total of 954 female youth who had experienced IPV in the past 12 months were included in the analysis, 681 from the 2015/16 TDHS and 273 from the 2022 TDHS. The median age was 21 years (IQR: 20,23) in the 2015/16 and 22 years (IQR: 20,23) in the 2022, with most participants aged 20-24, 510 (74.9%) in 2015/16 and 202 (74.0%) in 2022. Majority resided in rural areas 514 (75.5%) in 2015/16 and 212 (77.7%) in 2022, and were from Lake zone. Most were either married or cohabiting 541 (79.5%) in 2015/16 and 223 (81.7%) in 2022. had primary education 469 (68.9%) in 2015/16 and 161 (59.0%) in 2022 and belonged to the poor wealth index 322 (47.3%) in 2015/16 and 127 (46.7%) in 2022. While more than half were working in both surveys, there was a notable decline from 522 (74.9%) in 2015/16 to 140 (51.1%) in 2022. The median number of children was one, with nearly half reporting two or more children 332 (46.0%) in 2015/16 and 127 (46.5%) in 2022. In terms of empowerment, over 407 (75.8%) participated in at least one household decision in 2015/16, though this dropped to 131 (61.0%) in 2022. Justification of wife beating was common in both surveys 499 (73.2%) in 2015/16 and 179 (65.6%) in 2022, as was fear of partners 439 (64.4%) in 2015/16 and 170 (62.2%) in 2022. A smaller proportion reported experiencing severe violence, with a decline from 274 (40.2%) in 2015/16 and 69 (25.3%) in 2022. Witnessing interparental violence was comparable to those who had not across both surveys. Most had partners with primary education 347 (64.1%) in 2015/16 and 124 (57.9%) in 2022 and who did not consume alcohol 481 (70.6%) in 2015/16 and 214 (78.3%) in 2022 (Tables 2 and 3).

**Table 2.** Socio-demographic characteristics of Respondents in TDHS 2015/16 and 2022 (N= 954)

| Characteristic | Survey years: n (%) |  |
| --- | --- | --- |
|  | TDHS 2015/16 | TDHS 2022 |
|  | n = 681 | n = 273 |
| **Respondent's age (years)** |  |  |
| 15-19 | 171 (25.1) | 71 (26.0) |
| 20-24 | 510 (74.9) | 202 (74.0) |
| Median (IQR) | 21 (20,23) | 22 (20,23) |
| **Spousal age difference <sup>a b</sup>** |  |  |
| Wife is older | 2 (0.4) | 2 (1.0) |
| Wife is same age | 5 (0.9) | 5 (2.1) |
| Wife is 1-9 years younger | 410 (75.8) | 151 (70.6) |
| Wife is 10 or more years younger | 124 (22.9) | 56 (26.3) |
| **Residence** |  |  |
| Rural | 514 (75.5) | 212 (77.7) |
| Urban | 167 (24.5) | 61 (22.3) |
| **Zone** |  |  |
| Lake zone | 292 (42.9) | 127 (46.5) |
| Western | 127 (18.6) | 33 (12.1) |
| Eastern | 78 (11.4) | 29 (10.5) |
| Central | 51 (7.4) | 22 (8.0) |
| South West Highland | 45 (6.6) | 27 (9.9) |
| Northern | 31 (4.6) | 21 (7.9) |
| Southern Highland | 28 (4.2) | 7 (2.7) |
| Southern zone | 26 (3.8) | 5 (1.7) |
| Zanzibar | 3 (0.5) | 2 (0.7) |
| **Marital status at the time of interview** |  |  |
| Married/cohabiting | 541 (79.5) | 223 (81.7) |
| Widowed/divorced/separated | 140 (20.5) | 50 (18.3) |
| **Education level** |  |  |
| No education | 105 (15.5) | 65 (23.9) |
| Primary | 469 (68.9) | 161 (59.0) |
| Secondary/ higher | 107 (15.6) | 47 (17.1) |
| **Working status** |  |  |
| Not working | 159 (23.3) | 133 (48.8) |
| Working | 522 (76.7) | 140 (51.2) |
| **Wealth index** |  |  |
| Poor | 322 (47.3) | 127 (46.7) |
| Middle | 135 (19.7) | 52 (18.9) |
| Rich | 224 (33.0) | 94 (34.4) |
| **Number of living children** |  |  |
| None | 107 (15.7) | 64 (23.5) |
| 1 | 261 (38.3) | 82 (30.0) |
| 2+ | 332 (46.0) | 127 (46.5) |
| Median (IQR) | 1 (1,2) | 1 (1,2) |

IQR = Interquartile range, n = frequency, % is percentage, <sup>a</sup>2015/16 n==541; <sup>b</sup>2022 = 214

**Table 3.**
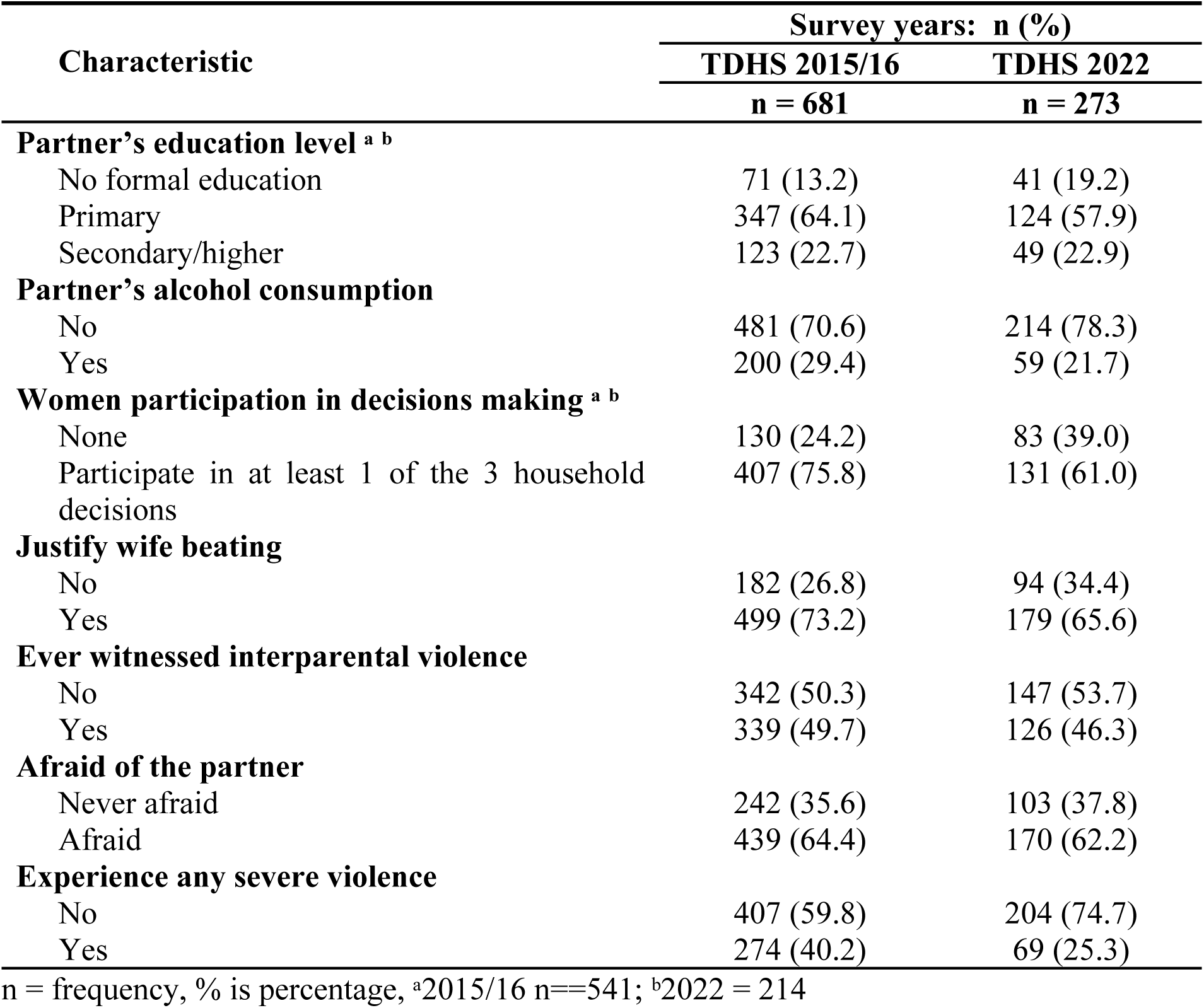
Partner-related characteristics, violence characteristics, and women’s empowerment indicators (N = 954)

### Prevalence of help-seeking behavior among female youth who experienced Intimate Partner Violence

Overall prevalence of help-seeking behavior among female youths who experienced Intimate Partner Violence in the past 12 months preceding the surveys decline significantly from 52.0% in 2015/16 to 34.1% in 2022 (p-value < 0.001) (Fig 2). In most regions the prevalence of help-seeking behavior declined except Manyara, Mwanza and Shinyanga. In 2015/16, Dodoma had the highest prevalence of help-seeking behavior (74.3%) followed by Singida (72.3%) and Arusha (71.9%). Regions with highest decline in help-seeking behavior are Singida, Arusha, Rukwa, Tanga and Dodoma (Fig 3)

**Fig 2.**
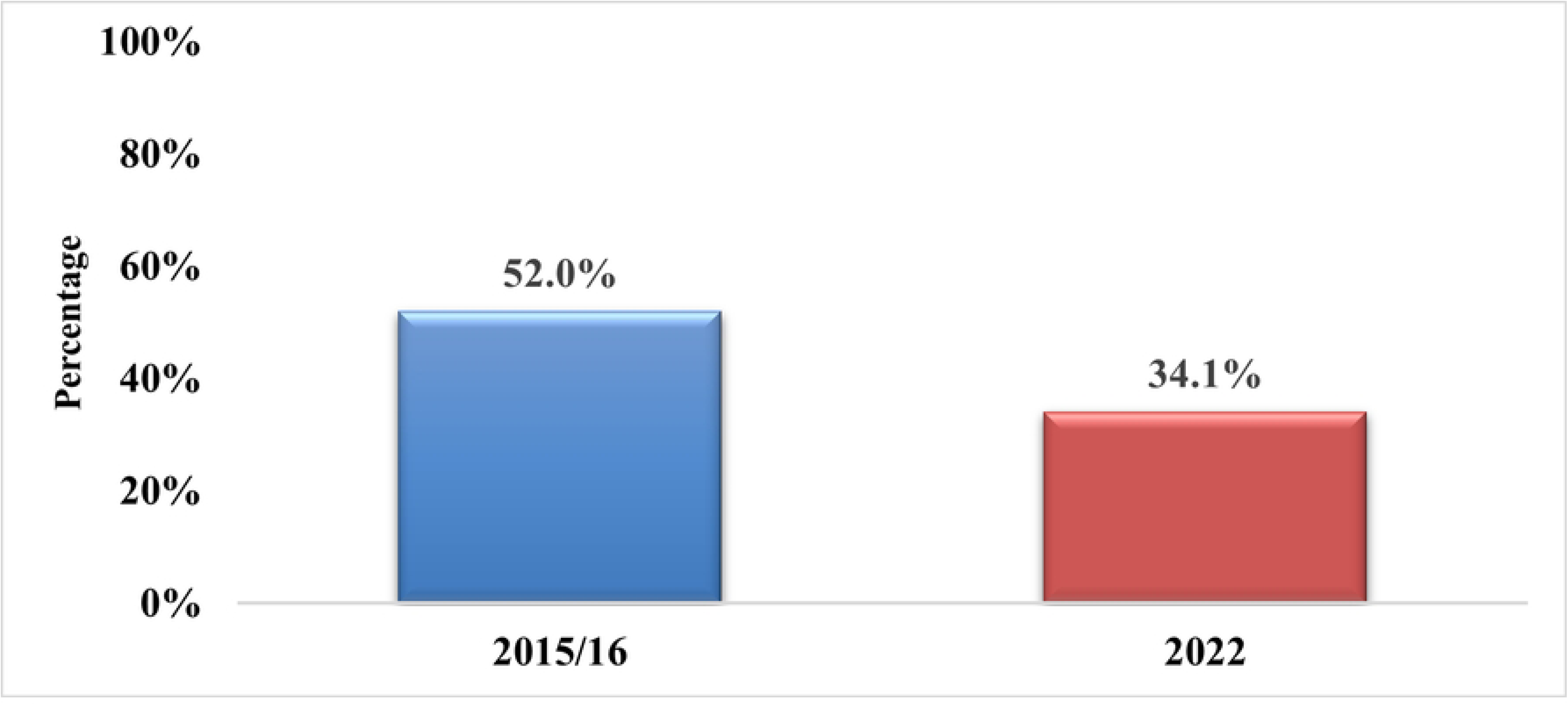
**Overall prevalence of help-seeking behavior by survey years**

**Fig 3.**
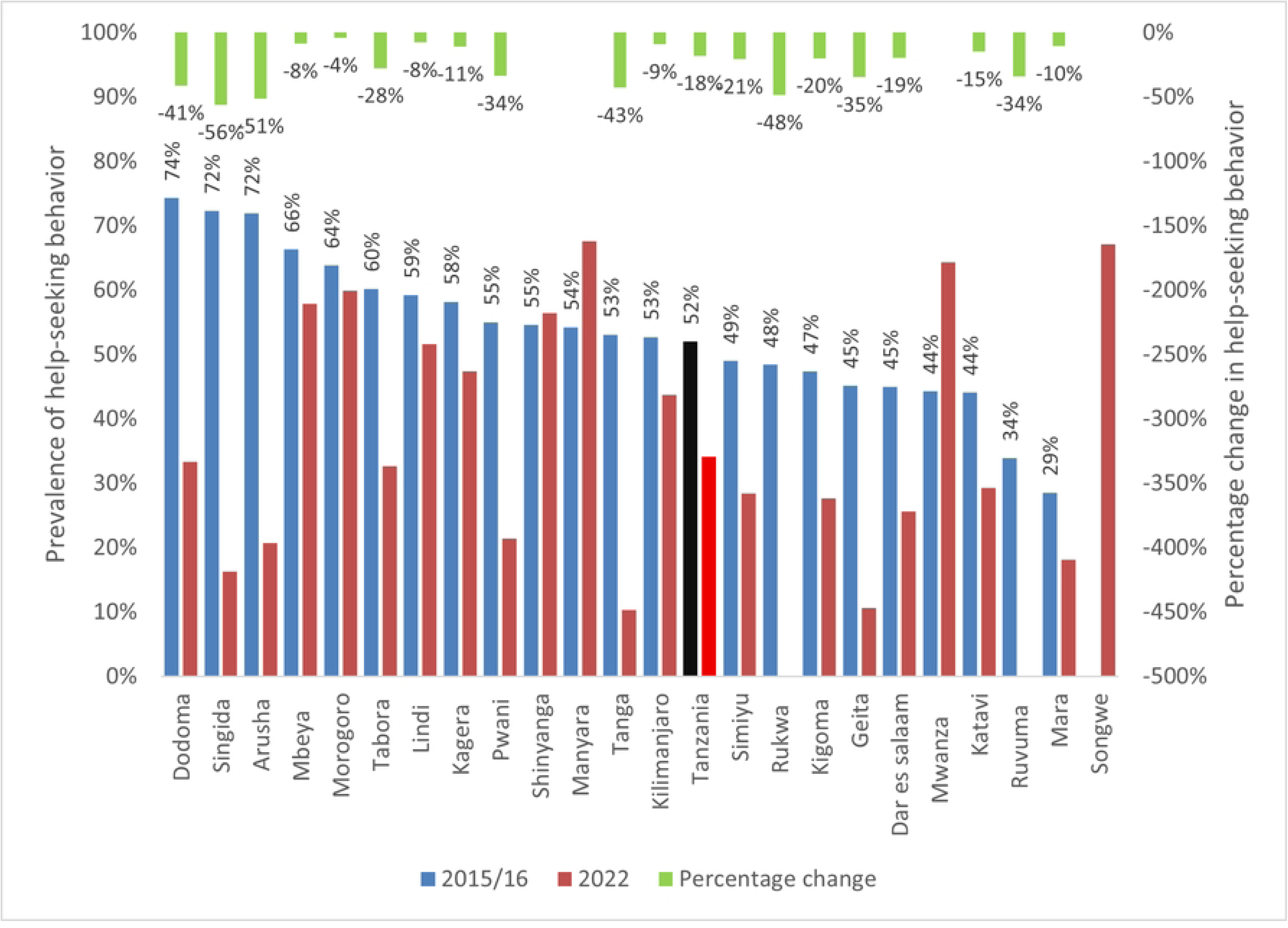
**Prevalence of help-seeking behavior and their percentage change by regions**

### Sources of help used by female youths who experienced IPV

In both surveys, the majority of female youths who reported seeking help after experiencing IPV did so from their own family (56.0% in 2015/16 and 41.4% in 2022) or from their husband’s/partner’s family (53.2% in 2015/16 and 45.8% in 2022). The least utilized sources of help were lawyers (1.5% in 2015/16 and 0% in 2022), medical personnel (0.5% in 2015/16 and 0% 2022) and social work organization (0.1% in 2015/16 and 1.2% in 2022 (Fig 4).

**Fig 4.**
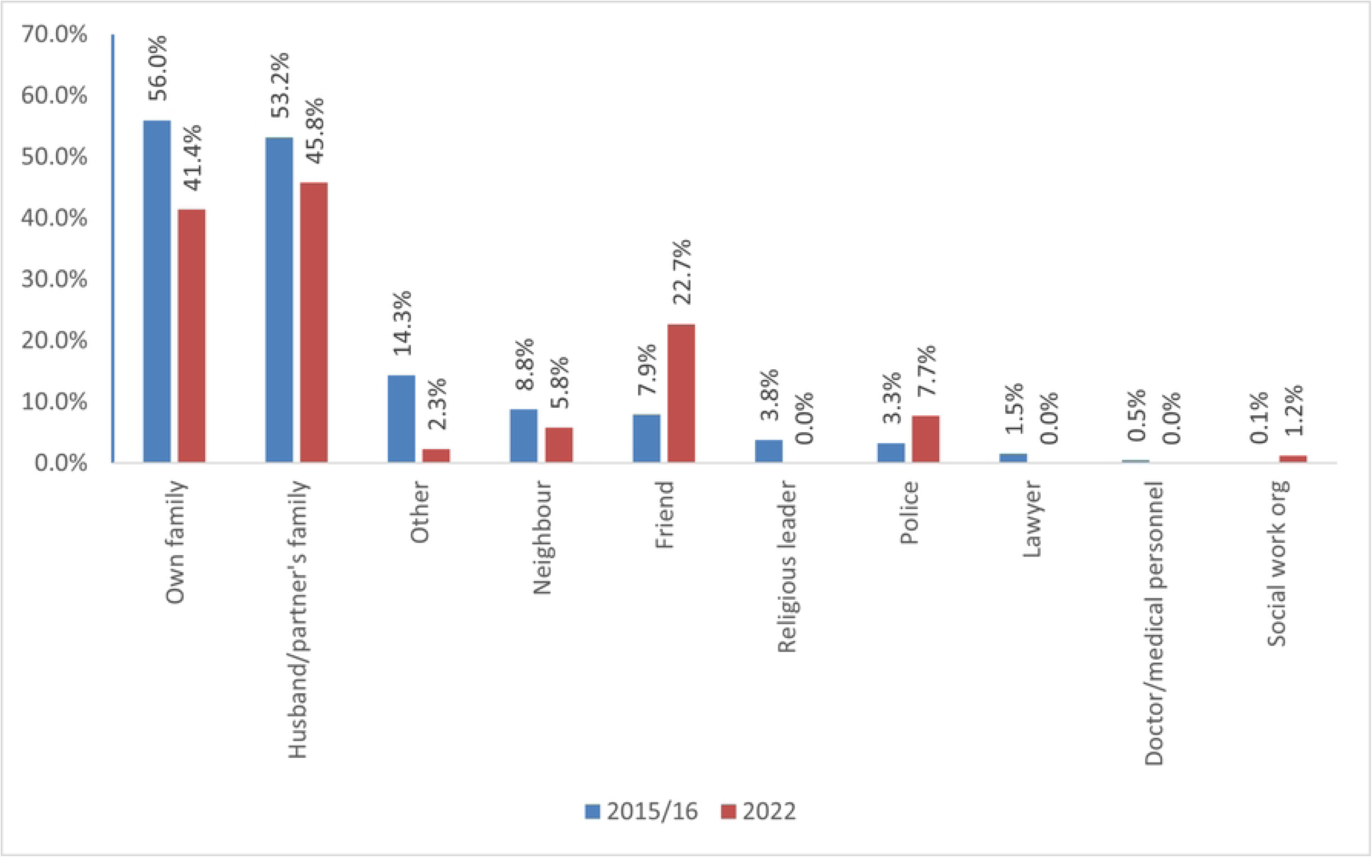
**Sources of help used by female youths who experienced IPV**

### Factors associated with help-seeking behavior among female youths who experienced IPV

After adjusting for other factors, in the 2022 TDHS, youth who were working had 82% higher prevalence of help-seeking behavior (aPR= 1.82; 95%CI: 1.15-2.88) compared to those who were not working. Similarly, youth in the rich wealth index had 76% higher prevalence of help-seeking behavior (aPR= 1.76; 95%CI: 1.05-2.97) compared to those in the poor wealth index. Spousal age difference was also significant factor significantly associated with help-seeking. Compared to youth who were older than their spouse, those who were one to nine years younger had 81% lower prevalence of help-seeking behavior (aPR = 0.19; 95%CI: 0.08-0.45), while those who were ten or more years younger had 82% lower prevalence (aPR= 0.18; 95%CI: 0.07-0.48) (Table 4).

**Table 4:** Factors associated with help-seeking behavior among female youths who experienced IPV.

| Variable | cPR (95%CI) | p-value | aPR (95%CI) | p-value |
| --- | --- | --- | --- | --- |
| **Respondent's age (years)** |  |  |  |  |
| 15-19 | 1 |  |  |  |
| 20-24 | 1.18 (0.67-2.07) | 0.576 |  |  |
| **Spousal age difference** |  |  |  |  |
| Wife is older | 1 |  | 1 |  |
| Wife is same age | 0.43(0.14-1.35) | 0.149 | 0.38(0.12-1.25) | 0.110 |
| Wife is 1-9 years younger | 0.30(0.21-0.42) | <0.001 | 0.19(0.08-0.45) | <0.001 |
| Wife is 10 or more years younger | 0.25(0.15-0.42) | <0.001 | 0.18(0.07-0.48) | 0.001 |
| **Zone** |  |  |  |  |
| Western | 1 |  |  |  |
| Southern Highland | 0.21 (0.03-1.78) | 0.153 |  |  |
| Northern | 0.59 (0.16-2.17) | 0.424 |  |  |
| Central | 1.12 (0.45-2.77) | 0.805 |  |  |
| Zanzibar | 0.96 (0.30-3.04) | 0.939 |  |  |
| Southern zone | 1.07 (0.29-3.92) | 0.918 |  |  |
| South West Highland | 1.43 (0.62-3.29) | 0.406 |  |  |
| Eastern | 1.30 (0.53-3.21) | 0.562 |  |  |
| Lake zone | 1.13 (0.52-2.46) | 0.758 |  |  |
| **Residence** |  |  |  |  |
| Urban | 1 |  |  |  |
| Rural | 0.63 (0.35-1.12) | 0.114 |  |  |

**Table 4: Factors associated with help-seeking behavior among female youths who** **experienced IPV (continuation)**
| Variable | cPR (95%CI) | p-value | aPR (95%CI) | p-value |
| --- | --- | --- | --- | --- |
| **Marital status** |  |  |  |  |
| Married/cohabiting | 1 |  |  |  |
| Widowed/divorced/separated | 2.09(1.29-3.37) | 0.003 | Omitted |  |
| **Working status** |  |  |  |  |
| Not working | 1 |  | 1 |  |
| Working | 1.72(1.06-2.81) | 0.029 | 1.82(1.15-2.88) | 0.011 |
| **Wealth index** |  |  |  |  |
| Poor | 1 |  | 1 |  |
| Middle | 0.82 (0.44-1.56) | 0.552 | 0.67(0.28-1.61) | 0.375 |
| Rich | 1.87(1.13-3.09) | 0.014 | 1.76(1.05-2.97) | 0.032 |
| **Severe violence experience** |  |  |  |  |
| No | 1 |  |  |  |
| Yes | 1.32(0.83-2.12) | 0.242 |  |  |
| **Participate in decision-making** |  |  |  |  |
| None | 1 |  | 1 |  |
| Participate in at least 1 of the 3 household decisions | 1.99(1.11-3.56) | 0.020 | 1.53(0.86-2.72) | 0.146 |
| **Afraid of the partner** |  |  |  |  |
| Never afraid | 1 |  |  |  |
| Afraid | 1.14 (0.60-2.18) | 0.689 |  |  |

**Table 4: Factors associated with help-seeking behavior among female youths who** **experienced IPV (continuation)**
| Variable | cPR (95%CI) | p-value | aPR (95%CI) | p-value |
| --- | --- | --- | --- | --- |
| **Witnessed interparental violence** |  |  |  |  |
| No | 1 |  | 1 |  |
| Yes | 0.63(0.40-0.99) | 0.048 | 0.77(0.49-1.21) | 0.264 |
| **Justify wife beating** |  |  |  |  |
| No | 1 |  | 1 |  |
| Yes | 0.69(0.41-1.16) | 0.161 | 0.81(0.48-1.37) | 0.438 |
| **Number of living children** |  |  |  |  |
| None | 1 |  |  |  |
| 1 | 0.73 (0.37-1.46) | 0.375 |  |  |
| 2+ | 0.75 (0.37-1.50) | 0.413 |  |  |
| **Respondent's education level** |  |  |  |  |
| No education | 1 |  |  |  |
| Primary | 1.24 (0.61-2.49) | 0.550 |  |  |
| Secondary/ higher | 1.50 (0.71-3.19) | 0.288 |  |  |
| **Partner's education level** |  |  |  |  |
| No formal education | 1 |  |  |  |
| Primary | 0.74 (0.35-1.55) | 0.424 |  |  |
| Secondary/higher | 1.06 (0.47-2.36) | 0.893 |  |  |
| **Partner's takes alcohol** |  |  |  |  |
| No | 1 |  |  |  |
| Yes | 0.91 (0.55-1.50) | 0.705 |  |  |

cPR= Crude prevalence ratio, aPR = Adjusted prevalence ratio, CI=Confidence interval

## Discussion

This study aimed to determine the prevalence of help-seeking behavior, describe sources of help used by and to determine the factors associated help-seeking behavior among female youth who experienced IPV in Tanzania in 2015/16 and 2022. The prevalence of help-seeking behavior significantly declined from 52% to 34.1% between 2015/16 and 2022 respectively. In both surveys, the most common sources of help were youth’s own family 56.0% in 2015/16 and 41.4% in 2022 and the partner’s family 53.2% in 2015/16 and 45.8% in 2022. In contrast, doctor/medical personnel and social welfare organizations were the least utilized sources, each reported by only 0.1% in 2015/16 and 1.2% in 2022, among youth who sought help. Help-seeking behavior was significantly associated with working, belonging to the rich wealth index, and being younger than the spouse.

The prevalence of help-seeking behavior among youth in this study declined significantly from 52% to 34.1% between 2015/16 and 2022 respectively. These findings are lower than that reported prevalence among the general population of women of reproductive age 15-49 years in Tanzania, as reported in respective TDHS reports, 54% in 2015/16 and 38% in 2022 (12,13). This suggests a disproportionately higher burden of unmet help-seeking needs among youth compared to the general population. However, the prevalence observed in this study remains higher than that reported in Kenya, where only 22% of youth sought help in 2022 and the reported prevalence in India 15% (17). The lower prevalence among youth compared to the general population may reflect limited interventions which are age appropriate for the young people and possible nonfunctional adolescent-youth friendly clinics in the health care facilities. Additionally, stigma, fear of exposing their partners and lack of family support may hinder youths from seeking help. Similar results were reported by (18) as barriers for youths to seek help. Furthermore, the observed decline in help-seeking behavior among youth indicate a failure to meet the national target set by the 2017/18-2021/22 NPA-VAWC, which aimed for 65% of survivors to report IPV within 72 hours (11). This could be attributed to the persistent influence of social and cultural barriers, particularly stigma, fear of community judgement and rooted gender norms (19).

Additionally, findings from this study indicate a higher preference of informal sources of help such as youth’s own family or partner’s family over formal sources of help including medical personnel and social welfare organizations in both surveys. This finding aligns with a study conducted in Tanzania by (4), which similarly reported more use of informal sources over formal sources. Several factors may contribute to these findings including, cultural norms that prioritize family privacy and belief that IPV should be resolved within the household. Other reasons could be limited awareness of available GBV health services among youths, particularly those who are not civil servants, some youth may not know whom to approach regarding IPV within health facilities, stigma surrounding IPV especially among adolescents and limited operating hours of the One-Stop centers may also hinder access to care. Moreover, lack of trust in formal sources such the police who are often perceived as prioritizing monetary gain over justice and fear of damaging family ties by exposing their spouse may further discourage formal help-seeking. Similar challenges were reported in qualitative studies conducted in Mwanza, Dar es Salaam, Iringa and Mbeya (19–21).

In this study, it was revealed that being in the rich wealth index was associated with a higher prevalence of help-seeking behavior. Similar results were reported in a study conducted in Ethiopia, which found that wealth increases the likelihood of seeking help (23). A possible explanation is that individuals from wealthier households often have better access to information and greater awareness of their rights and available sources of help, which may increase their likelihood of seeking help. Additionally, the financial independence can enhance autonomy and decision to seek help. A study conducted in Mbeya and Dar es Salaam reported that ownership of capital assets facilitated help seeking behavior (24). However, these findings contrast with those from a study that analyzed DHS data across 24 Sub-Saharan African countries, which found that being in the richest wealth quintile was associated with a lower likelihood of help-seeking (8). This discrepancy may be due to contextual differences on the role of wealth in shaping help-seeking behavior across countries.

Furthermore, youth who were working demonstrated a higher help-seeking prevalence. This may be attributed by greater financial independence and autonomy, which can empower youths to act against abuse. Similar findings were reported in a study that analyzed DHS data across 24 Sub-Saharan African countries (8), which reported that working or employed women were more likely to seek help. In contrast to these findings, a study conducted in three cities in Kenya, found no significant association between help-seeking behavior and working status among youth (25). The higher prevalence of help-seeking among working and wealthier youth supports previous findings reported in qualitative study by (20), that a major barrier to reporting IPV was lack of independent financial means to support themselves.

## Strength and limitation of the study

The study utilized data from the Tanzania Demographic and Health Survey, which was conducted across all regions of the country. Therefore, the findings are nationally representative and can be generalized to youth who have been exposed to intimate partner violence. Furthermore, the study utilized data from the two rounds of the TDHS, the first conducted prior to the implementation of the 2017/18-2021/22 NPA-VAWC, and the second conducted at the end of its implementation. This allows for a comparison of help-seeking behavior among youth before and after the national plan.

This study has several limitations. First, help-seeking behavior was measured using self-reported data which may be subject to reporting bias. This could result in either underestimation or overestimation of the true prevalence. Second, the analysis did not include all possible variables known to influence help-seeking behavior such as social network characteristics, mental health characteristics and awareness of available sources of help (25,28) , as these were not captured in the TDHS dataset

## Conclusion and recommendation

This study found a significant decline in help-seeking behavior among female youths who experienced IPV, with informal sources remaining the most preferred sources over formal in both surveys. Help-seeking behavior was significantly associated with belonging to the rich wealth index, working and being younger than the spouse. These findings highlight the critical role of families in early responses to IPV, suggesting that interventions should be strengthened by engaging families in IPV awareness and response efforts so they can provide timely and proper support through early detection, emotional support and referral services. The Ministry of Community Development, Gender, Women and Special groups (MoCDGWSG) and its partners should address cultural norms that discourage seeking help outside the family through school and media-based awareness interventions that tackle stigma and promote positive help-seeking attitudes. Youth who are not in school should be empowered economically so as to reduce their dependency on intimate partners. Increased economic autonomy may enhance their ability and willingness to seek help in cases of violence, as financial reliance often limits options for escape and recovery. Future research, particularly qualitative research, should focus on exploring the barriers among youth who do not seek help after experiencing IPV.

## Data Availability

Data cannot be shared publicly without written consent of DHS. Data are available from DHS MEASURE Program Data Access http://www.dhsprogram.com for researchers who meet the criteria for access the data

## Acknowledgements

We are grateful to the Almighty God for his guidance and protection. Special thanks to the academic staff at the School of Public Health, department of Epidemiology and Applied Biostatistics at KCMC University for their support throughout this study; Dr James Ngocho, Dr Shewiyo, Dr. William Nkenguye, Mr. Yusuph Nyaki, Mr. Baraka Mosha, Dr. Joachim Kessy.

## Author Contribution

**Conceptualization:** Asteria Karungi Nyongoli, Martha Magili, Winfrida C. Mwita, Caroline Amour, Sia E. Msuya

**Data curation:** Asteria Karungi Nyongoli, Anthony Kavindi, Idd A. Muruke

**Methodology:** Asteria Karungi Nyongoli, Laura J. Shirima, Winfrida C. Mwita, Caroline Amour, Sia E. Msuya

**Formal analysis:** Asteria Karungi Nyongoli

**Writing original draft**: Asteria Karungi Nyongoli

**Writing- review and editing**: Asteria Karungi Nyongoli, Winfrida C. Mwita, Caroline Amour, Sia E. Msuya

